# Understanding the United States Black-White Life Expectancy Gap, 2007-2018

**DOI:** 10.1101/2025.01.06.25319966

**Authors:** Harry Wetzler

## Abstract

**Background:** Life expectancy is a critical measure of population health. In the U.S., Black Americans have historically experienced lower life expectancy than White Americans due to factors such as health insurance inequities, socioeconomic disparities, and systemic barriers. Though the Black-White life expectancy gap narrowed after 1990, it has fluctuated in recent years, influenced by socioeconomic changes and the COVID-19 pandemic.

**Objective:** This study examines how national-level differences in education and income contributed to the Black-White life expectancy gap in the United States from 2007 to 2018.

**Methods:** Data were analyzed from the National Health and Nutrition Examination Survey (2007–2018) and its Linked Mortality File. Using 3 survival models, this study assessed life expectancy at age 20 for Non-Hispanic Black (NHB) and Non-Hispanic White (NHW) populations. Covariates included education and income.

**Results:** The unadjusted life expectancy gap at age 20 between NHB and NHW individuals averaged 3 years. Adjusting for education using a flexible parametric survival model reduced the gap by 50%, while adjusting for income reduced the gap by 75%. When both factors were adjusted simultaneously, two survival models indicated that NHB life expectancy slightly exceeded NHW life expectancy. Income disparities persisted across educational levels, signifying unequal economic returns to education.

**Conclusions:** Addressing income disparities is essential for reducing racial inequities in life expectancy. Policies promoting both equitable education access and income equivalence are critical for achieving health equity and improving population health.

## INTRODUCTION

Life expectancy or mean survival time is a common measure of health. Black people have historically had a lower life expectancy than White people in the United States (US). In 1940 the difference between Black and White life expectancy at age 20 was 8.7 years. This difference declined to 4.9 years in 2000 and to 2.9 years in 2014 but increased to 3.3 years in 2018 with a further pandemic-related rise to 4.6 years in 2021.^1^ Hill and Artiga suggested that inequities in health insurance coverage and access to care as well as social and economic factors are underlying drivers of these disparities.^2^ Schwandt, et al., found that after 1990, Black American life expectancy increased more than White American life expectancy in all US areas, but improvements in lower-income areas had the greatest impact on narrowing the racial life expectancy gap.^3^ Life expectancy for both Black and White Americans plateaued or slightly declined after 2012, but this slowdown was most evident among Black Americans even prior to the COVID-19 pandemic. In a systematic analysis of life expectancy by county, race, and ethnicity in the US over the years 2000 to 2019, the Global Burden of Disease (GBD) US Health Disparities Collaborators concluded that disparities in life expectancy among racial–ethnic groups in the US are widespread and enduring. They advocated accessing local-level data to address the root causes of poor health and early death among disadvantaged groups in the US.^4^

Previous studies have found strong associations between education, income, and life expectancy. Chetty et al. used complete income tax files (1.4 billion records) from 1999 through 2014 combined with mortality data from the Social Security Administration to study the association between income and longevity.^5^ Higher income was associated with greater longevity across the income continuum. The difference in life expectancy at age 40 between the highest and lowest income percentiles was over 10 years for women and 15 years for men.

Sasson and Hayward examined educational differences in adult life expectancy by analyzing 4.7 million deaths recorded in the US National Vital Statistics System in 2010 and 2017.^6^ Life expectancy at age 25 declined among white women and men with a high school degree or less by over a year. However, life expectancy increased among college-educated individuals.

This study aimed to evaluate how national-level education and income disparities contributed to the Black-White life expectancy gap in the United States between 2007 and 2018.

## METHODS

### Data Source

The National Health and Nutrition Examination Survey (NHANES) is a series of nationally representative cohort surveys conducted in two-year cycles, with linkage to the National Death Index. This analysis included adults aged 20 years or older from 2007 to 2018 because US life tables with specific results for Non-Hispanic Black and White populations were first published in 2006 (data from NHANES 2006 cannot be separated from 2005). Survey data were weighted to represent the noninstitutionalized civilian US population. All data and analytical guidelines are freely accessible from the US Centers for Disease Control and Prevention’s National Center for Health Statistics.^7^ The NHANES protocol was approved by the National Center for Health Statistics Institutional Review Board, and all participants provided written informed consent. This study adheres to the Strengthening the Reporting of Observational Studies in Epidemiology (STROBE) reporting guidelines.^8^

### Covariates

Age, race and ethnicity, educational attainment, and income were collected during in-home interviews. Race and ethnicity were self-reported using the following categories: Non-Hispanic Black (NHB), Non-Hispanic White (NHW), Mexican American, Other Hispanic, and Other Race - Including Multi-Racial. Education was categorized as less than 9^th^ grade, 9th to 11th grade, high school graduate, some college, and college graduate or above. Nineteen participants with missing education data were excluded from all analyses. Income was measured by the ratio of family income to the federal poverty level, adjusted for household size and inflation.

Approximate quartiles were constructed for the continuous income data, although 27.1% of the weighted sample fell into the top-coded category. Missing income data were coded as a separate category to maintain sample size.

### Data Analysis

Weighted tabulations of education and income distributions were conducted for NHB and NHW samples. Proportional hazards assumptions were tested using Schoenfeld residuals before fitting Cox, Weibull parametric, and flexible parametric survival models. Flexible parametric models, which use restricted cubic splines to model baseline excess hazards, were applied with degrees of freedom.^9^ Time-dependent effects (non-proportional excess hazards) were included with 3 degrees of freedom. Each survival model included race-ethnicity and a variable representing the NHANES two-year cycle for unadjusted results. Adjusted results were obtained by sequentially adding education and income as individual variables, followed by including both together in the model. Survival probabilities were estimated for each year from age 20 to 110, and life expectancy was calculated by integrating the area under the survival curve.

Analyses were conducted using Stata software version 18.5 (StataCorp). Confidence intervals were not calculated due to the exploratory nature of the study and limitations of Stata’s flexible parametric survival application (stpm3), which does not support complex survey analysis.

## RESULTS

### Sample Characteristics

The final sample consisted of 20,694 adults, including 7,195 NHB participants and 13,499 NHW participants. Table 1 lists the distribution of education and income for NHB and NHW participants. NHB participants were more likely to have lower educational attainment and income levels compared to NHW participants. NHB participants had a higher percentage of missing income data.

**Table 1.** Demographic Characteristics of the Sample (percentages)

| <u>Education</u> | **2007-2018** |  |
| --- | --- | --- |
|  | <u>NHB</u> | <u>NHW</u> |
| Less Than 9th Grade | 3.6 | 2.3 |
| 9-11th Grade<br>(Includes 12th grade with no diploma) | 16.4 | 8.3 |
| High School Grad/GED or Equivalent | 27.1 | 23.2 |
| Some College or AA degree | 34.7 | 32.5 |
| College Graduate or above | 18.2 | 33.8 |
| Income |  |  |
| Quartile 1 | 39.8 | 20.1 |
| Quartile 2 | 22.8 | 21.8 |
| Quartile 3 | 15.9 | 22.1 |
| Quartile 4 | 11.2 | 29.9 |
| missing | 10.4 | 6.1 |

### Income Disparities by Education

Table 2 compares income distributions by education level and race-ethnicity for high school graduates and above. Within the same educational category, NHW participants consistently had higher incomes compared to NHB participants. For instance, among college graduates, 51.8% of NHW participants were in the highest income quartile compared to 28.7% of NHB participants. Similarly, 37.4% of NHW high school graduates were in the top two income quartiles compared to 18.4% of NHB high school graduates. However, when education distributions were examined within income categories, these differences were not observed.

**Table 2.**
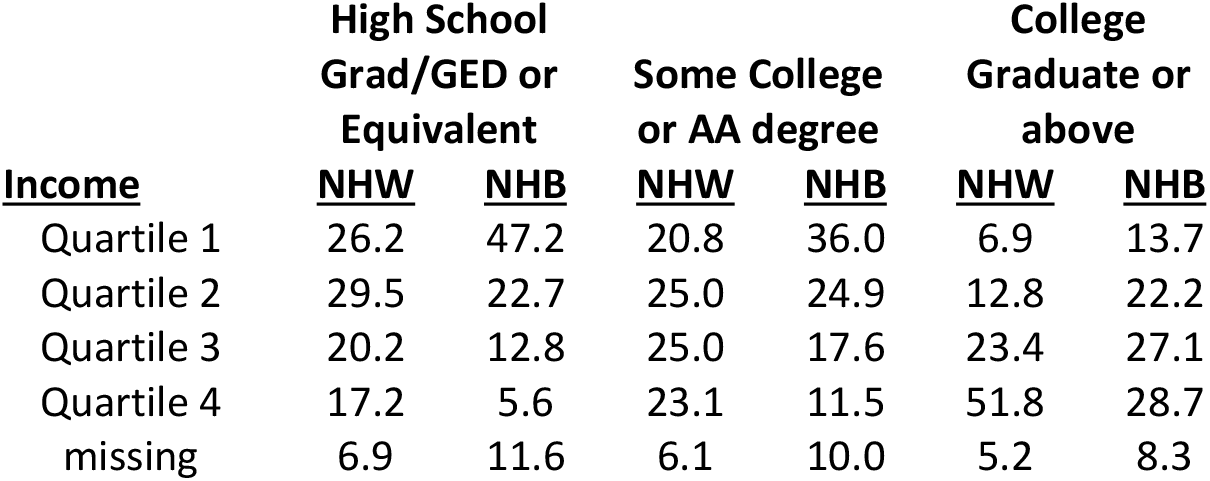
Income Distributions by Education and Race-Ethnicity (percentages)

### Life Expectancy Gaps

Table 3 shows the estimated gaps in life expectancy at age 20 between NHB and NHW participants, calculated using three different survival models. The unadjusted gaps were similar across models, averaging about three years, despite the proportional hazards assumptions failing for race-ethnicity, education, and income. Adjusting for education reduced the life expectancy gap by approximately half, while adjusting for income alone reduced the gap by 75%. When both factors were adjusted, NHB life expectancy slightly exceeded NHW life expectancy in two of the three survival models.

**Table 3.**
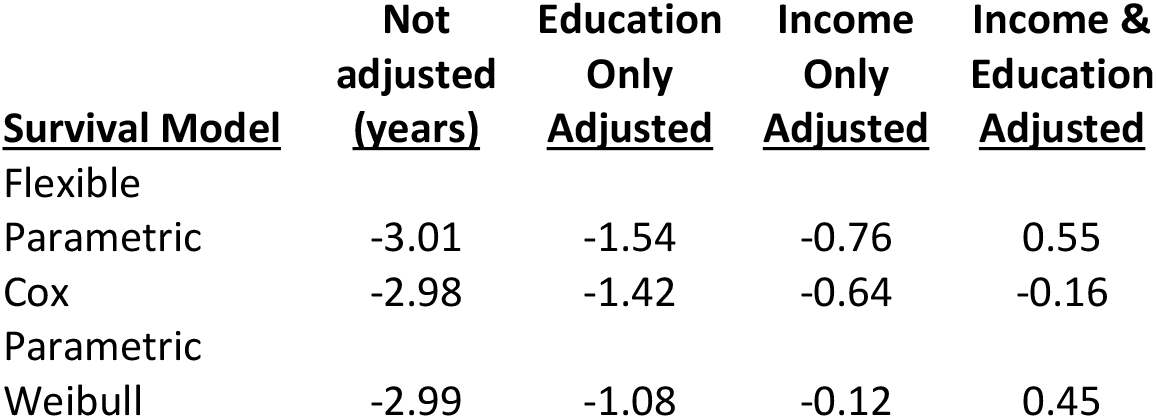
Estimated Black-White Life Expectancy Gaps at Age 20 (table values are NHB life expectancy minus NHW life expectancy)

## DISCUSSION

Adjusting for income eliminates three-fourths of the life expectancy gap between NHB and NHW individuals. By contrast, adjusting for education alone removes only about half of the gap. When both education and income are included, NHB life expectancy slightly exceeds that of NHW in two of the three survival models.

Income disparities persist even among individuals with comparable educational attainment. NHW individuals experience greater economic returns from higher education, a disparity well-documented in prior research. For instance, between 2000 and 2018, the Black-White racial wage gap widened across all educational levels, suggesting that higher education alone cannot resolve racial inequities in labor market outcomes.^10^ In 2023, the mean income of Black college graduates was $71,390, markedly lower than the $91,430 mean income of White college graduates.^11^

### Data Considerations

The accuracy of NHANES data in terms of race-ethnicity, education, income, and death linkage warrants scrutiny. Polubriaginof et al. found that data collected directly from respondents (as in NHANES) are the gold standard for capturing race and ethnicity, and Arias and colleagues reported high accuracy of race and ethnicity classification on death certificates for Black and White decedents.^12,13^

Educational data from NHANES, however, may include some misreporting. Foote and Warren found that 5–10% of individuals with bachelor’s degrees underreport their educational attainment, with higher rates of misreporting among Black respondents.^14^ For income, Moore, Stinson, and Welniak described systemic underreporting across U.S. government surveys.^15^

Although income underreporting is a recognized issue, its potential differential effects by race-ethnicity or education were not addressed. Notably, NHB participants in NHANES had a higher proportion of missing income data.

The quality of mortality linkages is another consideration. Studies, including those by Miller et al. and Lariscy, have identified slightly higher rates of missed death record links among Black individuals than White individuals, potentially introducing modest bias in mortality estimates.^16,17^

In this study, the average life expectancy gap between NHB and NHW individuals at age 20 was estimated to be 3 years during the period 2007–2018. However, NHANES data exclude individuals in institutional care and those who are incarcerated—populations that typically have shorter life expectancies.^18,19^ This exclusion is particularly significant given that Black Americans experience incarceration rates five times higher than White Americans.^20^ Including incarcerated individuals would disproportionately reduce NHB life expectancy and widen the NHB-NHW life expectancy gap. Notably, the overall U.S. average life expectancy gap at age 20 for the same period was 3.33 years.^1^ These results support the validity of the life expectancy estimates presented in this study.

### Methodological Considerations

This study used flexible parametric survival models to account for the non-proportional hazards observed for race-ethnicity, education, and income. No statistical model perfectly fits the data, but the general agreement across the three models provides confidence that the results are valid. Also, education and income were measured at a single point in time, and changes in these factors over the life course could lead to bias due to time-varying confounding.

### Implications and Future Directions

These results should be considered preliminary because NHANES has a relatively small sample size compared to the National Health Interview Survey (NHIS). To strengthen these findings, the next steps should involve analyzing additional NHANES cycles and examining NHIS data, which include mortality linkages dating back to 1986.

In 2016, Deaton emphasized the importance of education when discussing the Chetty study.^21^ He noted that the income-health link might stem from reverse causation where poor health reduces income, parental income shaping a child’s health and education, and education directly improving health and income through healthier habits, future planning, and better use of healthcare.

The finding that income strongly influences the NHB-NHW life expectancy gap aligns with the work of Wetzler and Cobb demonstrating that income inequalities have a greater impact on medical care utilization and expenditures than race or ethnicity among persons aged 0–64.^22^ While further research is needed to fully understand the mechanisms behind the income effect, these results underscore the urgent need for policies addressing income disparities to achieve health equity.

## CONCLUSION

Education and income play essential roles in improving life expectancy, yet their impacts differ in scope and immediacy. Education exerts a profound and enduring influence by shaping lifelong behaviors, expanding opportunities, and addressing broader social determinants of health. In contrast, income directly addresses immediate health needs and reduces disparities by improving access to resources, healthcare, and living conditions. Effective policies must address both factors simultaneously to reduce disparities and promote health equity.

## Data Availability

All data produced are available online at:
https://www.cdc.gov/nchs/nhanes/?CDC_AAref_Val=https://www.cdc.gov/nchs/nhanes/index.htm

